# A Replicable NeuroMark Template for Whole-Brain SPECT Reveals Data-Driven Perfusion Networks and Their Alterations in Schizophrenia

**DOI:** 10.64898/2026.04.08.26349985

**Authors:** Amritha Harikumar, Bradley Baker, Daniel Amen, David Keator, Vince D. Calhoun

## Abstract

Single photon emission computed tomography (SPECT) is a highly specialized imaging modality that enables measurement of regional cerebral perfusion and, in particular, resting cerebral blood flow (rCBF). Recent technological advances have improved SPECT quantification and reliability, making it increasingly useful for studying rCBF abnormalities and perfusion–network alterations in psychiatric and neurological disorders. To characterize large-scale functional organization in SPECT data, data-driven decomposition methods such as independent component analysis (ICA) have been used to extract covarying perfusion patterns that map onto interpretable brain networks. Blind ICA provides a data-driven approach to estimate these networks without strong prior assumptions. More recently, a hybrid approach that leverages spatial priors to guide a spatially constrained ICA (sc-ICA) have been used to fully automate the ICA analysis while also providing participant-specific network estimates. While this has been reliably demonstrated in fMRI with the NeuroMark template, there is currently no comparable SPECT template. A SPECT template would enable automatic estimation of functional SPECT networks with participant-specific expressions that correspond across participants and studies. The current study introduces a new replicable NeuroMark SPECT template for estimating canonical perfusion covariance patterns (networks). We first identify replicable SPECT networks using blind ICA applied to two large sample SPECT datasets. We then demonstrate the use of the resulting template by applying sc-ICA to an independent schizophrenia dataset. In sum, this work presents and shares the first NeuroMark SPECT template and demonstrating its utility in an independent cohort, providing a scalable and robust framework for network-based analyses.

## Introduction

SPECT is a highly specialized and clinically useful imaging modality for measuring cerebral perfusion, most commonly quantified as resting cerebral blood flow (rCBF) (Koyama et al., 1997). Recent technological advances have improved SPECT acquisition and quantification, increasing its utility as a robust measure of rCBF (Bouchareb et al., 2024; Harcourt, 2021; Koyama et al., 1997). Assessing rCBF with SPECT is valuable because perfusion abnormalities have been linked to a range of neurological and psychiatric conditions, including Alzheimer’s disease and schizophrenia (Boisvert et al., 2023).

To characterize covariance in perfusion patterns and rCBF abnormalities, sophisticated techniques such as independent component analysis (ICA) have been used to decompose functional brain data into independent components (Du et al., 2015). Functional neuroimaging-based templates such as NeuroMark have been applied using spatially constrained ICA (sc-ICA), which uses spatial priors and jointly maximizes 1) independence and 2) similarity of ICA components to the functional template (Du et al., 2020). While this approach has been reliably demonstrated in fMRI with the NeuroMark 1.0 template (Du et al., 2015, 2020), there currently is no comparable SPECT template for estimating functional brain networks from SPECT data. A replicable SPECT template would allow for the automation of ICA analyses in SPECT data, facilitating the investigation of relationships between network expression and clinical symptoms. Furthermore, the creation of a SPECT template would also enhance replicability, as estimated networks across studies correspond to the same template, facilitating comparison and replication of results, while still adapting to the individual data. The current study aims to address this gap in the current literature by introducing a novel NeuroMark SPECT template. Specifically, we performed blind ICA on two large-scale datasets to identify replicable components and generate the template. Here, we apply the template to estimate ICA from an independent schizophrenia dataset using sc-ICA. Results demonstrate reproducible patterns, particularly widespread decreased and increased patterns of co-modulation (CoM) in cognitive, subcortical, and sensorimotor network domains and subdomains which can be used in future studies to automatically estimate ICA while enhancing comparability across studies.

## Methods

We used three datasets in this study: two large datasets used to evaluate replicability of the components estimated from independent samples and for create the template, and a third, smaller dataset to demonstrate the application of the template to new data. SPECT data from patients with schizophrenia and healthy controls were obtained from the Amen Clinics (https://www.amenclinics.com/; see Tables 1a-1b below for demographic details, which are originally adapted from Harikumar et al., 2025). Blind ICA was run separately on sets of 5,001 (round 1), and 5,000 participants (round 2), randomly selected from a pool of 22,733 depressed individuals. One participant was excluded from round 2 after failing a manual quality control inspection (i.e. incorrect mask registration indicated by a zero-voxel value for that subject). A third group of SPECT images from 137 schizophrenia patients and 76 controls were analyzed by applying the new template using sc-ICA (Du et al., 2020b; Lin et al., 2010) with the multivariate objective optimization independent component analysis with reference (MOO-ICAR; Meng et al., 2023) algorithm in the Group ICA of fMRI Toolbox(GIFT; https://trendscenter.org/software/gift/). See Figure 1 for a graphical representation of the pipeline process. The NeuroMark SPECT template can be found here for download: http://trendscenter.org/data.

**Figure 1.**
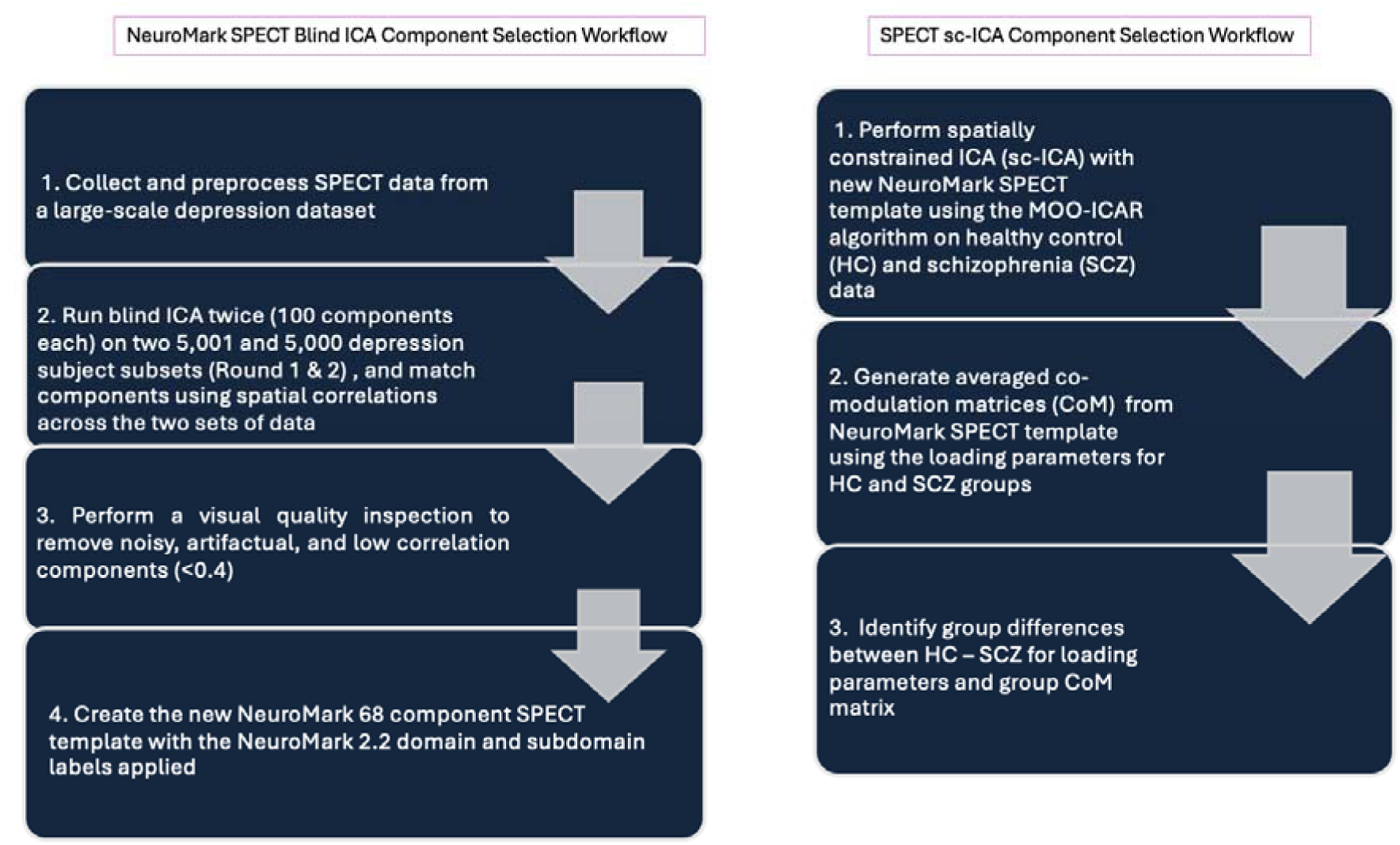
Comprehensive flowchart of the blind ICA component selection and sc-ICA template application processes.

**Table 1a.** Clinical Information of Patients and Racial Breakdowns.

Demographic information for patients with schizophrenia from the Amen Clinic dataset, with the site breakdown, average age and standard deviation, along with race classification information across sites.
| Amen Clinics Site # | Age (Mean \| SD) | Subject Breakdown | Gender | Categorization |
| --- | --- | --- | --- | --- |
| 1 | Between 21-25 \| 8.48 \| 2 subjects | 2M | N/A | 1 Hispanic/Latino, 1 African/American |
| 2 | 31.77 \| 11.08 \| 35 subjects | 17F/18M | 48%/51% | 1 Arab, 3 Asian, 19 Caucasian, 5 Hispanic/Latino, 1 Multiethnic, 1 Other, 5 Unknown |
| 3 | 38.5 \| 12.58 \| 12 subjects | 2F/10M | 16.6%/83.3% | 11 Caucasian, 1 Unknown |
| 4 | 33.14 \| 13.67 \| 27 subjects | 3F/24M | 11.1%/88.88% | 2 African/American, 23 Caucasian, 1 Hispanic/Latino, 1 Unknown |
| 5 | 37.66 \| 16.46 \| 12 subjects | 6F/6M | 50%/50% | 2 Asian, 8 Caucasian, 1 Native American, 1 Other |
| 6 | 25.77 \| 2.90 \| 9 subjects | 6F/3M | 66.6%/33.3% | 6 Caucasian, 1 Hispanic/Latino, 1 Other, 1 Unknown |
| 7 | 34.10 \| 15.32 \| 19 subjects | 4F/15M | 21.05%/78.94% | 1 African, 1 Asian, 15 Caucasian, 2 Unknown |
| 8 | 32.54 \| 11.08 \| 11 subjects | 6F/5M | 54.54%/45.45% | 2 Asian, 6 Caucasian, 1 Hispanic/Latino, 1 Other, 1 Unknown |
| 9 | 30.16 \| 10.22 \| 6 subjects | 2F/4M | 33.33%/66.66% | 4 Caucasian, 1 Multiethnic, 1 Unknown |
| 10 | Between 66-70 \| N/A (just 1 subject) | 1F | N/A | 1 Caucasian |
| **11** | Between 30-34 \| N/A (just 1 subject) | 1F | N/A | 1 Caucasian |
| **12** | Between 41-45 \| 22.62 \| 2 subjects | 1F/1M | 50%/50% | 2 Caucasian |
| **Totals** | 137 patients | 35% F/ 64% M | 39% F/ 60.1% M | 0.73% Arab / 5.8% Asian / 70.07% Caucasian / 6.6% Hispanic/Latino / 1.46% Multiethnic/ 2.92% Other / 8.7% Unknown / 2.19% African American / 0.73% Native American / 0.73% African |

**Table 1b.** Clinical Information of Controls and Racial Breakdowns.

Demographic information for healthy controls from the Amen Clinic dataset, with the site breakdown, average age and standard deviation, along with race classification information across sites. 12 sites were included in this study. Healthy controls were all from Site 2.
| Amen Clinics Site # | Age (Mean \| SD) | Subject Breakdown | Gender | Categorization |
| --- | --- | --- | --- | --- |
| **2** | Between 36-40 \| N/A \| 1 subject | 1M | N/A | African American |
| **2** | 48.71 \| 6.92 \| 7 subjects | 3F/4M | 42.85%/57.14%/ | Arab/Middle Eastern |
| **2** | 29.28 \| 6.92 \| 7 subjects | 4F/3M | 57.14%/42.85%/ | Asian |
| **2** | 43.74 \| 17.94 \| 55 subjects | 32F/23M | 58.18%/41.81% | Caucasian |
| **2** | 35.33 \| 9.266 \| 6 subjects | 3F/3M | 50%/50% | Hispanic/Latino |
| **Totals** | 76 controls | 55% F/ 44% M | 52% F/ 47% M | 1.32% African American/ 9.21% Arab / 9.21% Asian / 72.37% Caucasian / 7.89% Hispanic |

### Preprocessing Steps

Each patient participated in a SPECT brain scan acquired during rest across fourteen clinical imaging sites. SPECT scans were acquired using Picker (Philips) Prism XP 3000 triple-headed gamma cameras (Picker Int. Inc., Ohio Nuclear Medicine Division, Bedford Hills, OH, USA) with low energy high resolution fan beam collimators. For each procedure, an age- and weight-appropriate dose of 99mTc–hexamethylpropyleneamine oxime (HMPAO) was administered intravenously at rest. For the resting scans, patients were injected while they sat in a dimly lit room with their eyes open. Patients were scanned for approximately 30 minutes after injection. Data acquisition yielded 120 images per scan with each image separated by three degrees, spanning 360 degrees. A low pass filter was applied with a high cutoff and Chang attenuation correction performed (L.-T. Chang, 1978; W. Chang et al., 1984). The resulting reconstructed image matrices were 128×128×78 with voxel sizes of 2.5 mm^3^.

Next, images were aligned to the Montreal Neurological Institute (MNI) space with the Advanced Normalization Tools (ANTs version 2.2.0; Avants et al., 2011; RRID:SCR_004757) using a SPECT template, resulting in an image matrix size of 79×96×68 with isotropic voxel sizes of 2.0 mm^3^. SPECT images were scaled to the within-scan maximum voxel and noise outside of the brain was removed using a threshold of 50% of the maximum intensity, prior to registration. Next the mask images were aligned to the MNI space. The transformation was applied to the un-masked images. A brain mask derived from the MNI 152 (Fonov et al., 2011) template was used to remove noise outside the brain from the un-thresholded images for use in the statistical models. Registered SPECT scans were visually checked for the absence of severe functional abnormalities or artifacts and proper registration to the MNI space.

### Blind ICA Approach

Initial analysis was performed using the TReNDS/ARCTIC high performance computing cluster (https://arctic.gsu.edu/; see acknowledgment section for more information). For the blind ICA approach on the first two depression datasets, a standard Infomax algorithm (Bell & Sejnowski, 1994) was used with 100 components estimated for each dataset. After performing blind ICA, the 100 components underwent a multi-step quality control process. First, the Group ICA of fMRI toolbox (GIFT; http://trendscenter.org/software/gift) autolabeller tool (https://github.com/esalman/autolabeller) was utilized to identify a preliminary set of functional network categorizations. Afterwards, each component was visually inspected using the MRIcroGL package (https://www.nitrc.org/projects/mricrogl). After removing any artifactual components (e.g. components that were noisy, white matter) or those that had between-run spatial correlations < 0.4, 68 components remained. These were then labeled based on their similarity to the NeuroMark 2.2 template labels (Jensen et al., 2024; Figure 2) for each component.

**Figure 2.**
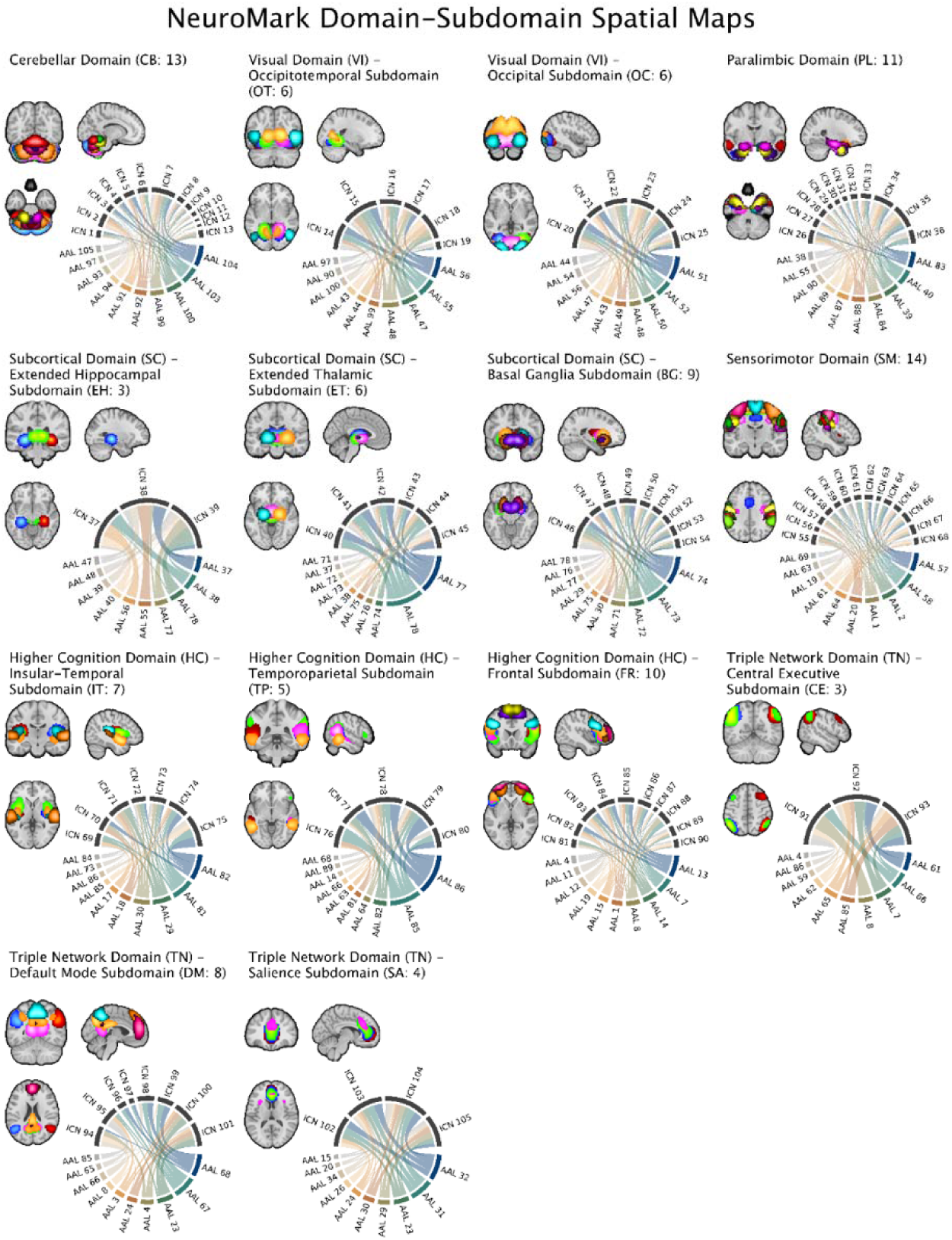
A visual depiction of the domains and subdomains with the corresponding components graphically and visually represented from NeuroMark 2.2 as adapted from Jensen et al., 2024.

### sc-ICA Approach

To demonstrate the efficacy of the NeuroMark SPECT template in application, we utilized GIFT to initially run the blind ICAs and sc-ICA. Afterwards, statistical analyses were run with GIFT using MATLAB R2020b to estimate participant-specific spatial maps using the new template as a reference. A total of 213 participants were used, and 68 components were estimated using the new NeuroMark SPECT template. Subsequently, the average co-modulation matrices (CoMs) were computed for both the control and patient groups, and a group difference CoM was calculated between these. Group differences in component expression across the 68 components were also evaluated.

## Results

### Blind ICA Results: Component Pairings

Below, we graphically display all 68 matched components on a spatial map (Figure 3) for round 1 (left) and round 2 (right). As expected, the maps highlight similarities in components across the separate Blind ICA analyses.

**Figure 3.**
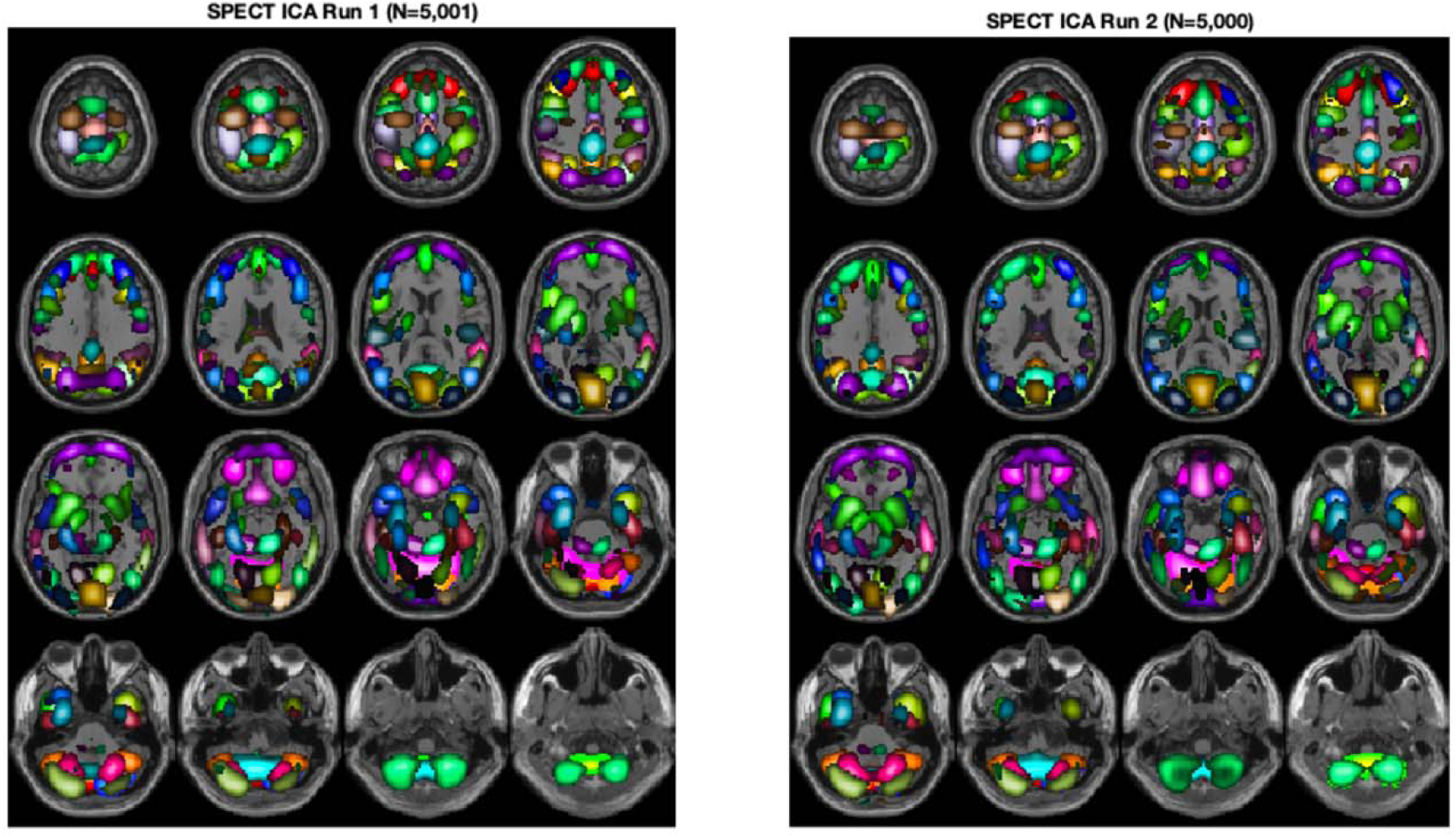
A graphical representation of the 68 matched components for Round 1 (left) and Round 2 (right) from the Blind ICA analyses. The high spatial correlation and strong visual correspondence across runs supports their stability and inclusion in the final NeuroMark SPECT template.

### Blind ICA Results: Co-modulation Matrix

A symmetric CoM matrix (Kotoski, 2026; Kotoski et al., 2024) was generated for each participant (component as the outer product of the loading parameters for each participant *i*) to understand individualized functional network connectivity for each participant, and verify component stability and reproducibility. The CoM matrix was computed for all 5,001 from th first dataset, and subsequently averaged across all participants (Figure 4). The CoM matrix can be used for comparing groups or participant variables, and was used after running the sc-ICA with the new NeuroMark SPECT template to examine group differences between schizophrenia (SZ) vs. healthy controls.

**Figure 4.**
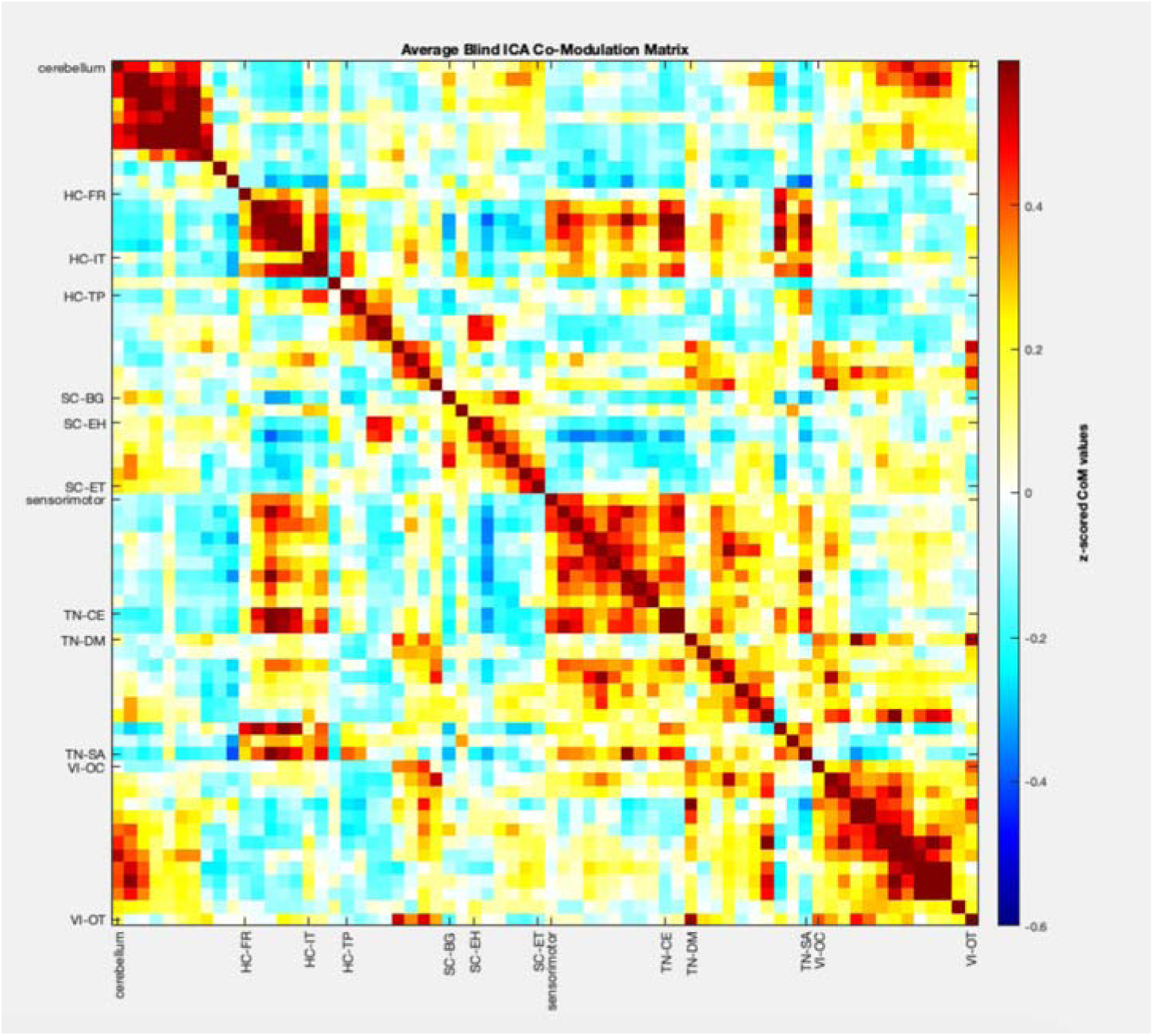
Average CoM matrix from the first run of blind ICA utilized to create the NeuroMark SPECT template. Here, the 68 components are organized by the respective NeuroMark 2.2 domains and subdomains listed as the following: cerebellar (CB), higher cognition-frontal (HC-FR), higher cognition – insular-temporal (HC-IT), higher cognition-temporoparietal subdomain (HC-TP),subcortical-basal ganglia (SC-BG), subcortical – extende hippocampal (SC-EH), subcortical – extended thalamic (SC-ET), sensorimotor (SM), triple network – central executive (TN-CE), triple network – default mode (TN-DM), triple network-salience (TN-SA), visual-occipital (VI-OC), and visual-occipitotemporal (VI-OT) subdomains. Results show highly modular organization, with both short (within domain) and long (between domain) range covariation.

### Application to Patient vs. Control Cohort: Loading Parameter Results and Group Differences

To evaluate group differences in the expression of SPECT networks, we first computed a two-sample t-test (control – patient) to investigate group differences across the *loading parameters* (e.g. how well components were expressed across the 68 components) using the new SPECT template. After correcting for multiple comparisons using the false discovery rate (FDR) correction (Benjamini & Hochberg, 1995), 23 out of the 68 components were found to be statistically significant between controls-patients, and subsequently visualized and represented in the spatial maps and bar plot below (Figures 5 and 6). These results show SPECT signal differences in the cross-network co-modulation patient and control brains based on the CoM matrices across cognitive, visual, cerebellar, and sensorimotor areas between healthy patients and controls. Specifically, increased cerebellar average loading values for patients > controls suggest patterns like what we noted above. Overall, patients demonstrated higher loadings for cerebellar, SC-EH, SC-ET, and VI-OC subdomains, compared to increased widespread domain and subdomain loadings in controls across the HC-FR, HC-TP, and HC-IT regions. Particularly important to note is the increased temporal lobe loading in Component 23, which could be indicative of clinical symptoms in schizophrenia (e.g. auditory hallucinations) in patients. Taken together, these results suggest that dysfunction could be segregated to certain disconnected networks in schizophrenia, centered around subcortical, cerebellar, and triple networks (Friston, 1998; Harikumar et al., 2023; Menon, 2011).

**Figure 5.**
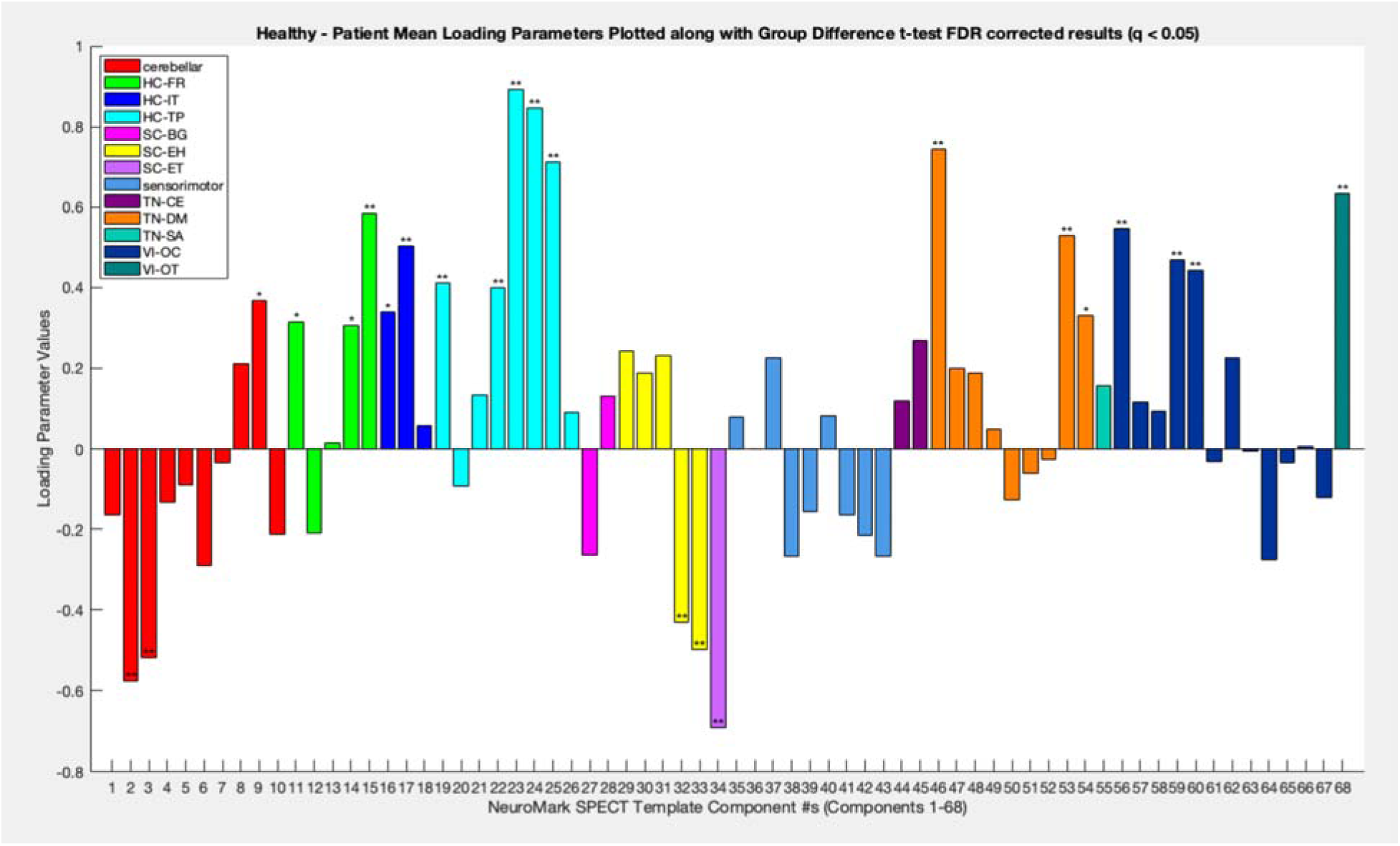
Shows a bar plot with mean loading components plotted after performing a t-test between healthy controls and patients. 23 significant components (5 as SZ > HC, 18 as HC > SZ) after performing FDR correction *(q < 0.05*; q< 0.01**)* showed that increased loading parameter functional covariance was noted across components pertaining to the HC-FR, HC-IT, HC-TP, TN-DM, VI-OC, and VI-OT subdomains for controls. In contrast, increased loading parameter expression in patients was noted in cerebellar, SC-EH and SC-ET subdomains as expected.

**Figure 6.**
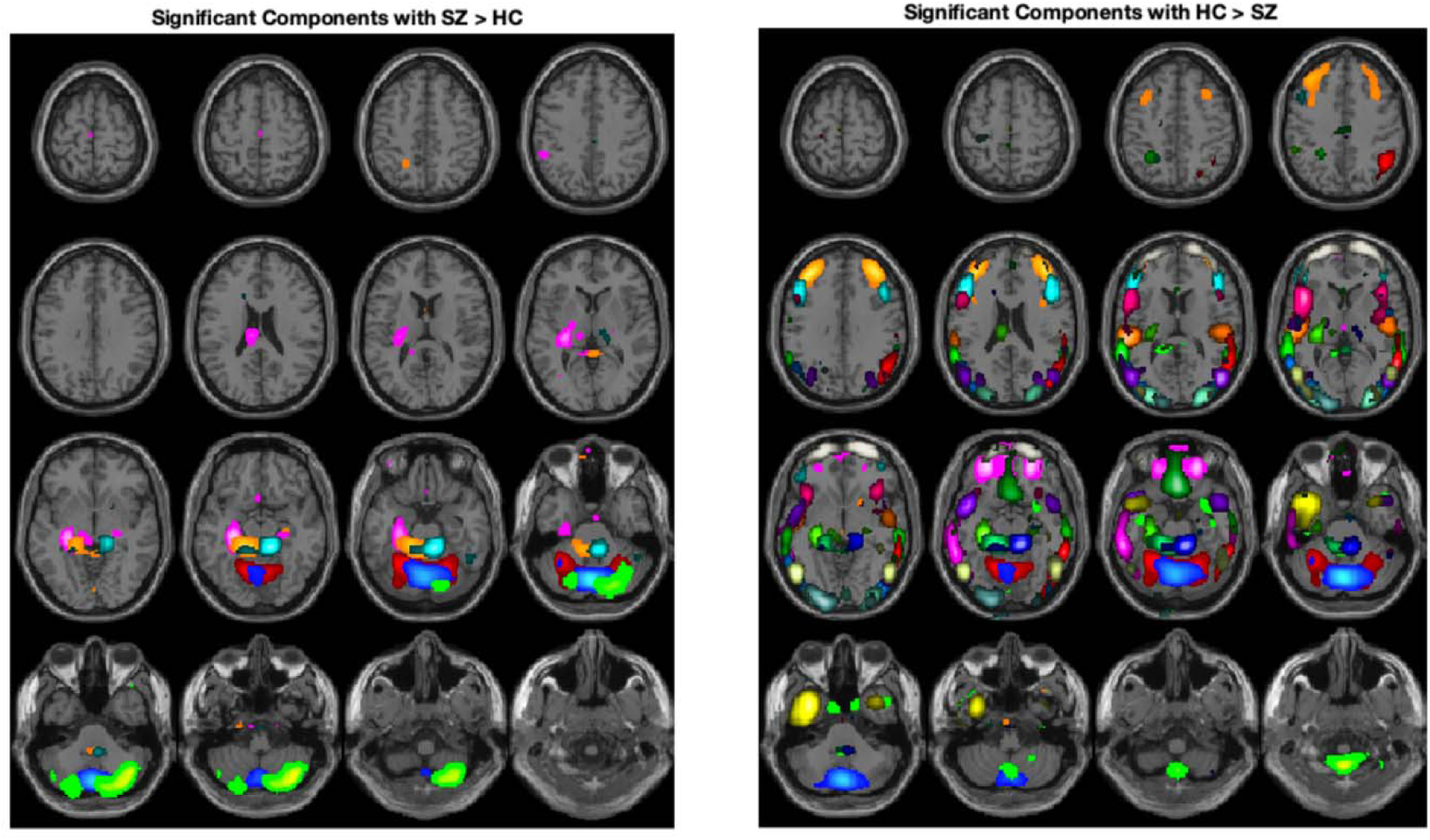
Components showing significant loading parameter differences, On the left the five components with higher loading parameter expression in patients; on the right are shown the 18 components with higher loading parameter expression in controls. Patients showed more cerebellar/subcortical activity, compared to controls wh showed more HC subdomain activity.

### Application to Patient vs. Control Cohort: CoM Matrix Results and Group Differences

Next, group averaged CoM matrices were calculated across the NeuroMark SPECT template components. Matrices were calculated for controls and patients separately as well as for a group difference between controls – patients. For controls (Figure 7; left), an expected pattern of widespread CoM, particularly increased/decreased CoM across the domains and subdomains persisted. Here, decreased average CoM was notably found in cerebellar, subcortical, frontal, and thalamic regions. Consequently, increased average CoM was found in triple network regions and default mode network, subcortical hippocampal, default mode network, and visual systems. This suggests a more distributed pattern of covariance in controls, where the cognitive, executive, and visual systems seem to function cohesively as stable networks. The patient group (Figure 8; right) however, showed different CoM patterns, indicating a possibility of atypical functional reorganization in patients. Notably, overall concentrated, increased CoM patterns in certain domains and subdomains, rather than a wide spread of increased/decreased CoM as seen in controls was noted in patients. These patterns of increased CoM, particularly in regions such as the cerebellar-SC-EH, SC-BG, and SC-ET subdomains suggest that patients showed disrupted functioning across various brain regions related to cerebello-thalamocortical circuits, which may be associated with clinical symptomatology. Particularly, increased CoM in subcortical and cognitive control regions may be indicative of executive functioning deficits in patients.

**Figures 7 and 8.**
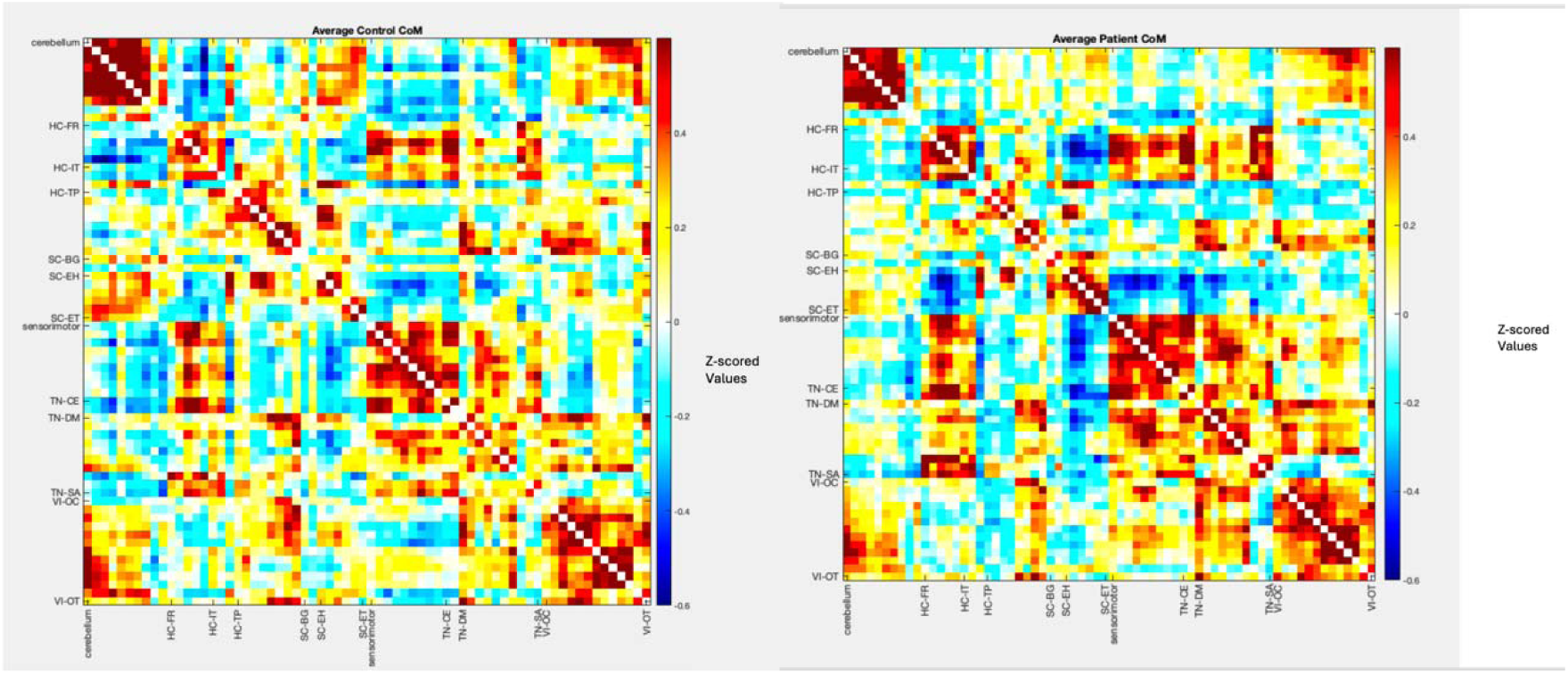
Average co-modulation for the controls (left) and patients (right). Both patients and controls showed the expected modular structure in the CoM matrix, with controls showing a more widespread CoM pattern across the different domains and subdomains. In contrast, patients showed increased CoM in areas such as the SC-ET, SC-EH, SC-BG, cerebellar, visual, and HC domains and subdomains.

**Figure 8a-8b.**
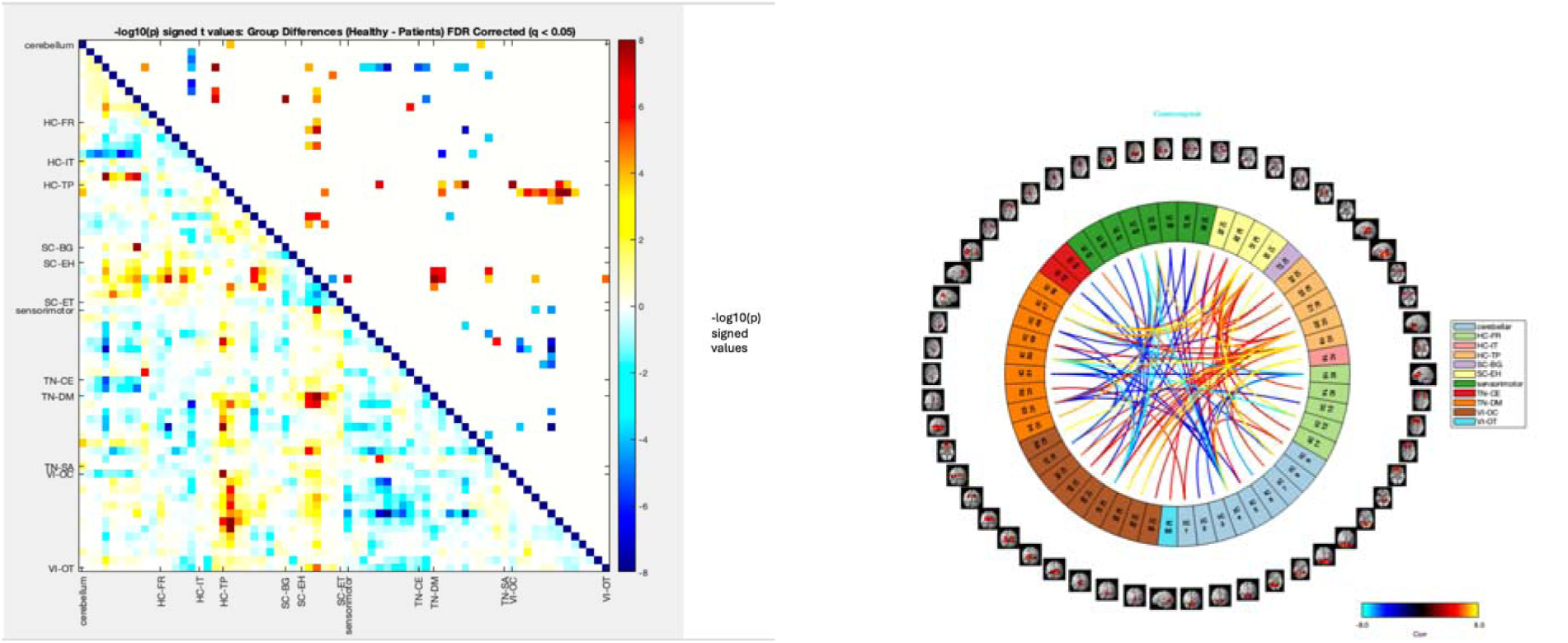
Group differences between control-patients averaged CoM matrices plotted with all values (Figure 8a, left) on the lower left triangle, and non-significant thresholded -log10p signed values zeroed out on the upper right half of the triangle. Here, warmer colors indicate regions with increased CoM between domains and subdomains. Patterns show that patients displayed increased CoM in cerebellar, SC-EH, SC-BG, HC-TP, and VI-OC subdomains; controls showed more widespread increased/decreased CoM across HC, TN, SC, and sensorimotor domains/subdomains. Figure 8b (right) demonstrates the upper triangle -logt10(p) significant results plotted as a connectogram, visually demonstrating the components with significant CoM results.

Finally, an examination of controls– SZ group differences show that patients have stronger CoM across the visual, higher cognition, triple network, and subcortical networks. Figure 8a (left) shows the -log10(p) * signed t-value plot for a two-sample t-test on the CoM matrix, with the upper triangle showing FDR corrected results. Figure 8b (right) shows the corresponding connectogram showing significant results. Results also show a decreased CoM pattern across subcortical, cognitive, paralimbic, and visual areas in controls. This suggests that patients may have disrupted cerebello-thalamo-cortical and visual circuitry, which may relate to positive and negative symptom domains such as auditory and visual hallucinations. The results in Figure 8a-8b also replicate the patterns seen for patients and controls; namely, increased CoM in visual and cerebello-thalamocortical circuits in patients, increased/decreased CoM across all domains and subdomains for controls.

## Discussion

These results support the utility of the newly created SPECT NeuroMark template, and it ability to delineate CoM patterns within controls and patients, as well across group comparisons.

First, the components selected for the template were replicated across two independent sets of data (N≈5,000 each). The resulting networks were largely consistent with those found in resting fMRI studies. Some subcortical structures such as the caudate were less prominently represented, potentially reflecting modality-specific sensitivity limitations. The application of NeuroMark SPECT to the schizophrenia cohort revealed several interpretable patterns which may inform our understanding of cognition and behavior. CoM has been utilized in prior source based morphometry (SBM) studies to measure structural covariation (Kotoski, 2026), which found widespread reductions in co-modulation in schizophrenia, particularly within and between areas such as the DM, VI, and cognitive networks.

Additionally, we were able to find similar patterns of disruption in the CoM. Unlike fMRI-derived functional connectivity, SPECT-derived networks reflect spatial covariance in perfusion rather than temporal synchrony. This distinction is important when interpreting CoM results. Individuals with schizophrenia are known to experience disruptions broadly across the triple network and default mode networks (Wu et al., 2025), with evidence of altered structural abnormalities in schizophrenia across cognitive and default mode networks as well (Salgado-Pineda et al., 2011). These structural disruptions (Karlsgodt et al., 2010) could point to various emotion/face processing deficits in schizophrenia (Salgado-Pineda et al., 2011), as well as deficits in cognition and working memory (Karlsgodt et al., 2010). These disruptions could also point to a broad failure to disengage the DM in patients (Karlsgodt et al., 2010; Salgado-Pineda et al., 2011; Wu et al., 2025), which point to cognitive deficits and decreased attentional resources in their overall functioning.

We noted similar patterns in our study across similar networks, with previous findings indicating increased loadings in the same dataset across auditory, subcortical, and sensorimotor regions (Harikumar et al., 2025) which were connected to auditory hallucination symptomatology. Previous reviews and studies (Hare, 2021; Hare et al., 2018; Harikumar et al., 2023; Jensen, Calhoun, et al., 2024) indicate complex patterns of network dysconnectivity, including aberrant cerebello-thalamocortical circuitry and salience network disruptions in schizophrenia. Using another modality such as SPECT imaging may provide complementary information about perfusion-related activity relative to fMRI, or independently. The current study provides a novel framework and template for which future studies can investigate some of these patterns of dysconnectivity in greater detail. This SPECT template could also be further utilized to help understand clinical symptoms in schizophrenia.

We also note the relatively modest sample size in the schizophrenia cohort as a possible limitation. Further research is needed, particularly along the lines of investigating CoM across a larger, and more heterogeneous neuropsychiatric group to better understand these patterns. Additionally, correlations to specific positive and negative clinical symptoms in schizophrenia to the CoM is warranted to further understand brain-behavior relationships.

### Conclusions

To conclude, we presented the creation and demonstrated the utility of the first NeuroMark SPECT template. We identified key patterns of CoM differences in controls vs. SZ, as well as in controls and patients, and we identified significant component differences. Overall, results replicated a widespread, complex pattern of CoM as seen in previous studies (Kotoski, 2026), and warrant further investigation with large sample sizes. Future studies can correlate CoM to clinical symptoms, and utilize the new SPECT template to compare with prior fMRI studies, and integrate findings across imaging modalities.

## Data Availability

Data is restricted due to privacy/confidentiality for patients, but any requests can be directed to Dr. David Keator.

## Acknowledgments

We acknowledge the use of Advanced Research Computing Technology and Innovation Core (ARCTIC) resources at Georgia State University’s Research Solutions made available by the National Science Foundation Major Research Instrumentation (MRI) grant number CNS-1920024.

## Statement of Informed Consent/Approval

Integ Review IRB, Protocol: 004-Amen Clinics Inc., Determination: Exempt Category 4. Approval Date: 9/19/2014. Study Title: Retrospective Review of Clinical Cases in a Brain SPECT Imaging Database. Investigator: Daniel Amen, M.D. Integ Review IRB was acquired by Advarra IRB.

## Declaration of Generative AI and AI Assisted Technologies in the Writing Process

During the preparation of this work the primary author used Grammarly, Microsoft Copilot, and ChatGPT to check content for flow and clarity, verify simple summation totals for tables, create and streamline MATLAB code for efficiency, and proofread the work for grammar and spelling errors. After using this tool/service, the primary author reviewed and edited the content as needed and take(s) full responsibility for the content of the published article.

